# Correlation analysis of risk factors and GSI score of a medical team assisting Wuhan city during the epidemic of COVID-19 in China -A cohort study

**DOI:** 10.1101/2020.04.27.20070466

**Authors:** Cheng Wang, Jinlong Zhang, Zhaohui Lu, Jingquan Wang, Yunyun Fang, Yanlin Wang, Xia Chen, Na Hong, Xiaolei Jing

## Abstract

**Importance:** There are few studies on the psychological status of medical staff during the COVID-19 outbreak. This study is the first in the world about the psychological status of the medical team during the COVID-19 outbreak.

**Objective:** To study the correlation between risk factors and general symptom index (GSI) score of medical team members who support Wuhan against COVID-19.

**Design:** Cohort study.

**Setting:** Population-based.

**Participants:** Anhui Province sent a total of eight medical teams, including 1382 members, to support Hubei Province. We adopted a stratified sampling method and selected the fourth team sent by Anhui Provincial Hospital, with a total of 137 members as our subjects.

**Exposures:** Four main exposures were collected, including basic information, preparations before going to Wuhan, life issues and working issues after going to Wuhan.

**Main Outcomes and Measures:** The GSI score of SCL-90 scale was used to reflect the frequency and intensity of psychological symptoms. We made the hypothesis of this study before data collection.

**Results:** 110(80.29%) members completed the questionnaire, of which, 77(70.00%) female and 33(30.00%) male. When adjusted age, gender and covariates, DC, LCWT had a positive correlations with GSI score(β was10.17, 95%CI was 3.30 to 17.04 for DC, *P*=0.00<0.05; β was 11.55, 95%CI was 0.40 to 22.71 for LCWT, *P* =0.04<0.05;respectively), RBT had a negative positive correlation with GSI score (β was -28.09, 95%CI was -45.79 to -10.40, *P*=0.00<0.05), AoBI did not had a correlation with GSI score (β was 11.55, 95%CI was 0.40 to 22.71, *P*=0.16>0.05). When adjusted covariates, DC had a positive and RBT had a negative correlation with GSI score of female (β was 13.20, 95%CI was 4.55 to 21.85, *P*=0.00<0.05; β was -57.85, 95%CI was -94.52 to -21.18, *P*=0.00<0.05; respectively), but for male was not (*P*=0.59>0.05, *P*=0.08>0.05, respectively), LCWT and AoBI didn’t had correlation with GSI score between genders (*P*>0.05).

**Conclusions and Relevance:** Improving DC, RBT and decreasing LCWT can reduce the GSI score. AoBI didn’t affect the psychological status; male members have a more stable mood than female. Whether other countries medical team has the same result still needs further research.

**Key Points:** *Question:* What is the correlation between risk factors and general symptom index (GSI) score of medical team members who support Wuhan against COVID-19?

*Findings:* Dietary conditions (DC) had a positive and relationship between team (RBT) had a negative correlation with GSI score of female, but for male was not (*P*=0.59>0.05, *P*=0.08>0.05, respectively), lacking communication with teams (LCWT) and afraid of being infected (AoBI) didn’t had correlation with GSI score between genders, a significant difference.

*Meaning:* Improving DC, RBT and decreasing LCWT between team members can reduce the GSI score. Whether they are AoBI, didn’t affect the psychological status, male members have a more stable mood than female.

## 1. Introduction

COVID-19 was endemic in China at the end of 2019. Data as received by WHO from national authorities by 10:00 CEST, 23 April 2020, there were 2 544 792 confirmed cases and 175 694 deaths^1^, In this epidemic, China has quickly taken various effective measures. Now the epidemic is basically under effective control in China. These experiences are worth sharing with the world. One of the measures was to recruit medical personnel from various provinces and cities across the country to support the most severely affected province-Hubei, with a total of nearly 40,000 medical personnel. This type of disease has caused global panic, and medical staffs are no exception, they may also feel panic^2-4^. The current research mainly focuses on the psychological status of infected people^5-7^. Few people pay attention to the psychological symptoms of medical staff a study found that it is helpful to take effective interventions to meet their needs if the needs of nurses caring for COVID-19 patients could be perceived well, the main needs was health and security^8^. At the time of the COVID-19 epidemic outbreak in China, the heroic Chinese medical staffs were under the tremendous pressure of fighting the COVID-19, whether they had a poor psychological status? And what the risk factors were? So far, there was no research report and also no similar literature in other countries around the world when we search the database on line.

Symptom Checklist-90 (SCL-90) scale has been widely used in psychology research in the world. The author used this scale to evaluate ordinary people psychology status during COVID-19 pandemic in China, and found which also had a significant adverse socio-psychological influence on ordinary citizens^9^. It has good reliability and validity^10, 11^, and can reflect the mental and psychological status of the recipients from multiple dimensions. The GSI score of SCL-90 can reflect the frequency and intensity of psychological symptoms^12, 13^. Therefore, this study analyzed the GSI score of a medical team, we carried out systematic statistical analysis to find out the risk factors that affected the GSI score, in order to provide a theoretical basis for precise psychological intervention for medical staffs.

## 2. Methods

### 2.1 Design methods

We used a cohort study, using the single-blind method; the participants were not clear about the purpose.

### 2.2 Data collecting methods

We used Questionnaire star APP (https://www.wjx.cn/) to collect the data, the collected data set includes the basic information, preparations before going to Wuhan, life issues and working issues after going to Wuhan. The basic information included seven sub risk factors-age, working age, education background, only child of the family, marriage status, childbirth status and family relationships. There were five sub risk factors of preparations before going to Wuhan, including COVID-19 knowledge, infection prevention and control knowledge, confidence to complete the task, emergency aid experience and relationships with team (RBT). There were seven sub risk factors of life issues after going to Wuhan, including dietary conditions (DC), sleep quality (SQ), limit range of activities (LRoA), care about me (CAM), lavation conditions (LC), residential conditions (RC) and surrounding conditions (SC). There were nine sub risk factors of working issues after going to Wuhan, which were unfamiliar with medical records system (UWMRS), unfamiliar with working environment (UWWE), difficulty communicating with patients (DCWP), unfamiliar with workflow (UWW), lacking medical equipments (LME), long working hours (LWH), lacking communication with teams (LCWT), difficulty communicating with local hospital (DCWLH) and fear of being infected (AoBI). The GSI score of SCL-90 scale was used to reflect the frequency and intensity of psychological symptoms

### 2.3 Statistical methods

In this survey study we took a purposive approach with no sample estimation. Data were analyzed using the statistical packages R (R Foundation; http://www.r-project.org;version3.4.3) and EmpowerStats (http://www.empowerstats.com; X&Y Solutions Inc, Boston, MA). Multivariable logistic regression model, single factor logistic regression model, generalized estimated equation model, T-test and Pearson’s chi-square tests were used to analyze the data, estimated change (β) and 95% confidence interval(CI) were used to establish the demographic and clinic characteristic of the sample, results were considered statistically significant with *P*<0.05.

### 2.4 Ethical considerations

The participants of this study filled out the questionnaire anonymously. Our research was approved by the Ethics Committee of the First Affiliated Hospital of University of Science and Technology of China (Anhui Provincial Hospital).

## 3. Results

### 3.1 Flow chart of the study

Anhui Province sent a total of eight medical teams to support Hubei, with a total of 1,362 medical members, including a total of 274 in the fourth medical team, which were selected from the First Affiliated Hospital of Anhui Medical University and the First Affiliated Hospital of University of Science and Technology of China (Anhui Provincial Hospital). The first author of this study was one of the members of the latter medical constitution, so the object was the members of the latter as for the convenience of research. For details, see Fig1.

**Figure1.**
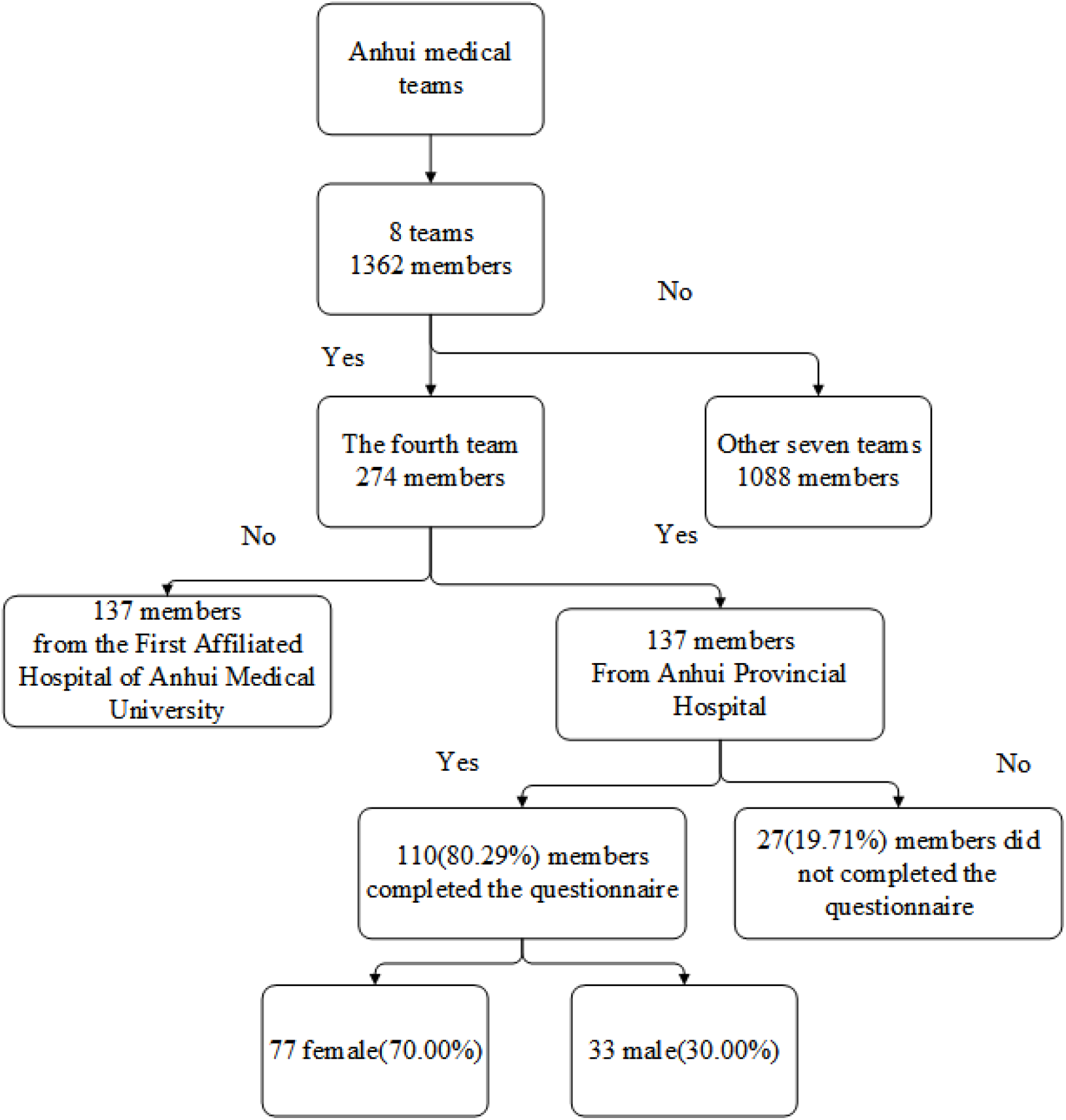
Flow chart of the study.

### 3.2 Demographic and risk factors of members

110(80.29%) members completed the questionnaire, of which, 77(70.00%) female and 33(30.00%) male. For basic information, 76.6% female and 69.7% male had a bachelor degree, the difference of education background ratio between the genders was statistically significant(X^2^=9.77, df=3, *P*=0.02<0.05). 41.6% female and 21.2% male unmarried, the marriage ratio between the genders was also statistically significant(X^2^=4.18, df=1, *P*=0.04<0.05). For preparations before going to Wuhan, the relationships with team (RBT) factor had a statistically significant between genders, 98.7% female and 90.9% male felt a good interpersonal relationship with the team(X^2^=4.00, df=1, *P*=0.04<0.05).For life issues after going to Wuhan, 96.1% female and 84.8% male felt care about me(CAM) from the team, and the differences also had a statistically significant (X^2^=4.34, df=1, *P*=0.04<0.05).The other factors of genders had no statistically significant(*P*>0.05).

### 3.3 Crude correlation associations of exposure risk factors and GSI score of the members

As seen in table2,we analyzed the exposure risk factors, the single factor analysis showed risk factors of basic information had no correlation with GSI score(*P*>0.05). For preparations before going to Wuhan, there were five risk factors, RBT had a negative correlation with GSI score, estimated change was -20.63, 95%CI was -39.65 to -1.60, the difference had a statistically significant(*P*=0.04<0.05). For life issues after going to Wuhan, there were seven risk factors, DC had a positive correlation with GSI score, estimated change was 11.08, 95% CI was 4.11 to 18.05, the difference had a statistically significant(*P*=0.00<0.05). For working issues after going to Wuhan, there were nine risk factors, LCWT and AoBI had a positive correlation with GSI score, the estimated change was 17.11 and 16.32 respectively, 95%CI was 6.32 to 27.90 and 5.08 to 27.57 respectively, the difference had a statistically significant (*P*=0.00<0.05,P=0.01<0.05,respectively).

**Table 1.** Demographic and risk factors of the sample: 77 women and 33 men

| Risk factors | Mean± SD/ N% |  | P Value |
| --- | --- | --- | --- |
|  | Women ( 77 ) | Men ( 33 ) |  |
| **Basic information** |  |  |  |
| Age(years) |  |  | 0.24 |
| 20-29 | 36.4 | 21.2 |  |
| 30-39 | 50.6 | 57.6 |  |
| >39 | 13.0 | 21.2 |  |
| Working age ( years ) |  |  | 0.21 |
| <5 | 24.7 | 15.2 |  |
| 6-10 | 33.8 | 39.4 |  |
| 10-20 | 32.5 | 30.3 |  |
| 21-30 | 9.1 | 9.1 |  |
| >30 | 0.0 | 6.1 |  |
| Education background |  |  | 0.02<sup>*</sup> |
| College | 11.7 | 0.0 |  |
| Bachelor | 76.6 | 69.7 |  |
| Master | 11.7 | 27.3 |  |
| Doctor | 0.0 | 3.0 |  |
| Only child of the Family |  |  | 0.22 |
| Yes | 19.5 | 30.3 |  |
| No | 80.5 | 69.7 |  |
| Marital Status |  |  | 0.04<sup>*</sup> |
| Married | 58.4 | 78.8 |  |
| Unmarried | 41.6 | 21.2 |  |
| Childbirth status |  |  | 0.17 |
| Yes | 55.8 | 69.7 |  |
| No | 44.2 | 30.3 |  |
| Family relationships |  |  | 0.86 |
| Good | 94.8 | 93.9 |  |
| Poor | 5.2 | 6.1 |  |
| **Preparations before going to Wuhan city** |  |  |  |
| Infection prevention and control knowledge |  |  | 0.17 |
| Familiar | 84.4 | 93.9 |  |
| Unfamiliar | 15.6 | 6.1 |  |
| COVID-19 knowledge |  |  | 0.47 |
| Familiar | 89.6 | 93.9 |  |
| Unfamiliar | 10.4 | 6.1 |  |
| Confidence to complete the task |  |  | 0.12 |
| Yes | 100.0 | 96.8 |  |
| No | 0.0 | 3.2 |  |
| *Emergency aid experience* |  |  | 0.97 |
| Yes | 42.9 | 42.4 |  |
| No | 57.1 | 57.6 |  |
| *RBT* |  |  | 0.04* |
| Good | 98.7 | 90.9 |  |
| Poor | 1.3 | 9.1 |  |
| **Life issues after going to Wuhan city** |  |  |  |
| *DC* |  |  | 0.97 |
| Good | 48.1 | 48.5 |  |
| Poor | 51.9 | 51.5 |  |
| *SQ* |  |  | 0.11 |
| Good | 35.1 | 51.5 |  |
| Poor | 64.9 | 48.5 |  |
| *LROA* |  |  | 0.45 |
| Adapted | 44.2 | 36.4 |  |
| Unadapted | 55.8 | 63.6 |  |
| *CAM* |  |  | 0.04* |
| Yes | 96.1 | 84.8 |  |
| No | 3.9 | 15.2 |  |
| *LC* |  |  | 0.53 |
| Good | 57.1 | 63.6 |  |
| Poor | 42.9 | 36.4 |  |
| *RC* |  |  | 0.62 |
| Good | 96.1 | 93.9 |  |
| Poor | 3.9 | 6.1 |  |
| *SEC* |  |  | 0.07 |
| Good | 90.9 | 78.8 |  |
| Poor | 9.1 | 21.2 |  |
| **Working issues after going to Wuhan city** |  |  |  |
| *UWMRS* |  |  | 0.92 |
| Yes | 77.9 | 78.8 |  |
| No | 22.1 | 21.2 |  |
| *UWWE* |  |  | 0.40 |
| Yes | 48.1 | 39.4 |  |
| No | 51.9 | 60.6 |  |
| *DCWP* |  |  | 0.93 |
| Yes | 59.7 | 60.6 |  |
| No | 40.3 | 39.4 |  |
| *UWW* |  |  | 0.28 |
| Yes | 50.6 | 39.4 |  |
| No | 49.4 | 60.6 |  |
| *LME* |  |  | 0.82 |
| Yes | 7.8 | 9.1 |  |
| No | 92.2 | 90.9 |  |
| *LWH* |  |  | 0.86 |
| Yes | 5.2 | 6.1 |  |
| No | 94.8 | 93.9 |  |
| *LCWT* |  |  | 0.32 |
| Yes | 14.3 | 6.1 |  |
| No | 85.7 | 87.9 |  |
| *DCWLH* |  |  | 0.20 |
| Yes | 5.2 | 12.1 |  |
| No | 94.8 | 90.3 |  |
| *AoBI* |  |  | 0.29 |
| Yes | 13.0 | 6.1 |  |
| No | 87.0 | 93.9 |  |
| ***SCL-90 Score*** |  |  |  |
| GSI | 1.20±0.24 | 1.17±0.16 | 0.56 |

Note: GSI=general symptom index, RBT= relationship between team, DC=dietary conditions, SQ=Sleep quality, LRoA=limit range of activities, CAM=care about me, LC=lavation conditions, RC=residential conditions, SEC= surrounding environmental conditions, UWMRS=unfamiliar with medical records system, UWWE=unfamiliar with working environment, DCWP=difficulty communicating with patients, UWW=unfamiliar with workflow, LME=lacking medical equipments, LWH=long working hours, LCWT=lacking communication with teams, DCWLH=difficulty communicating with local hospital, AoBI= afraid of being infected.

**Table2.**
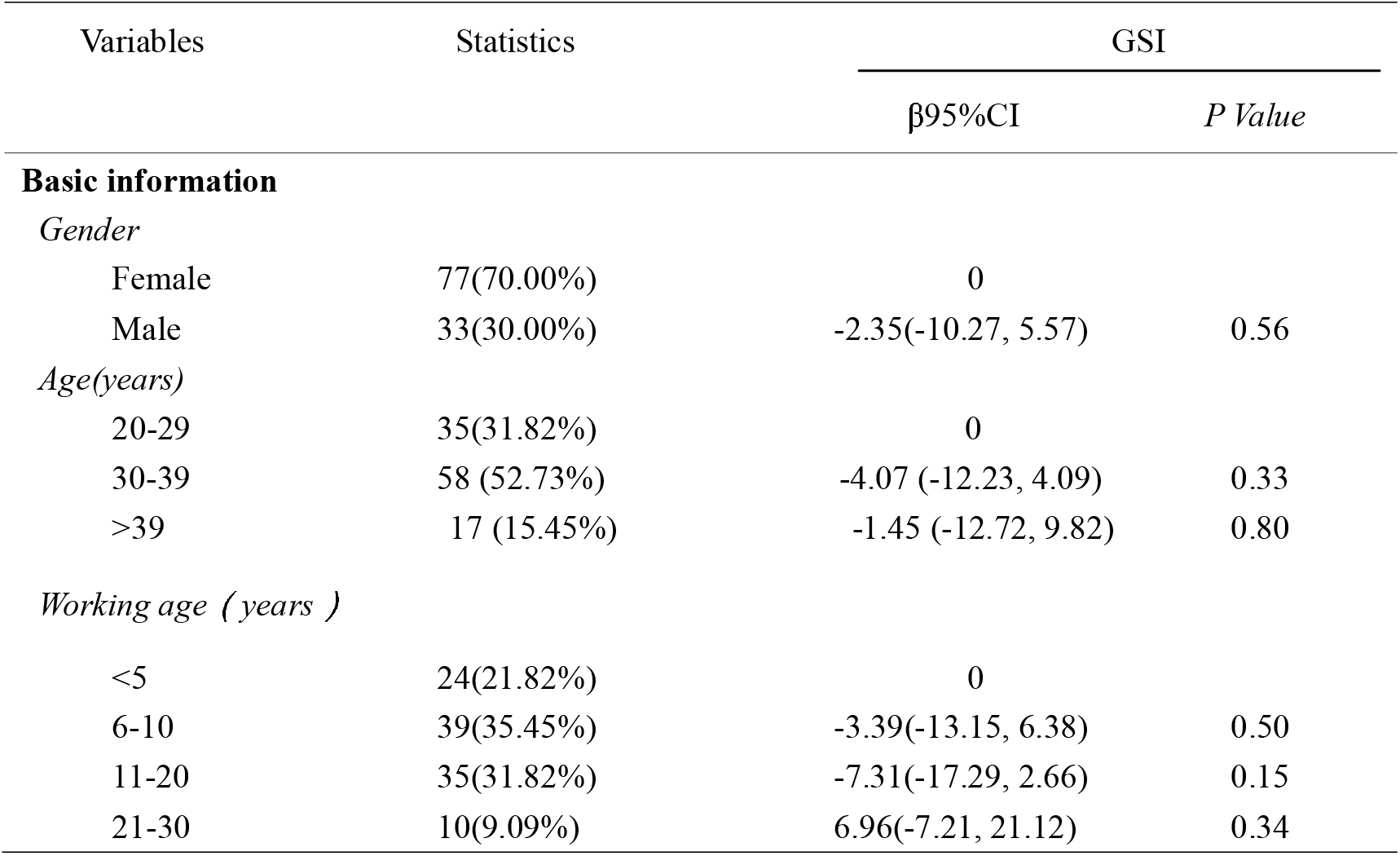
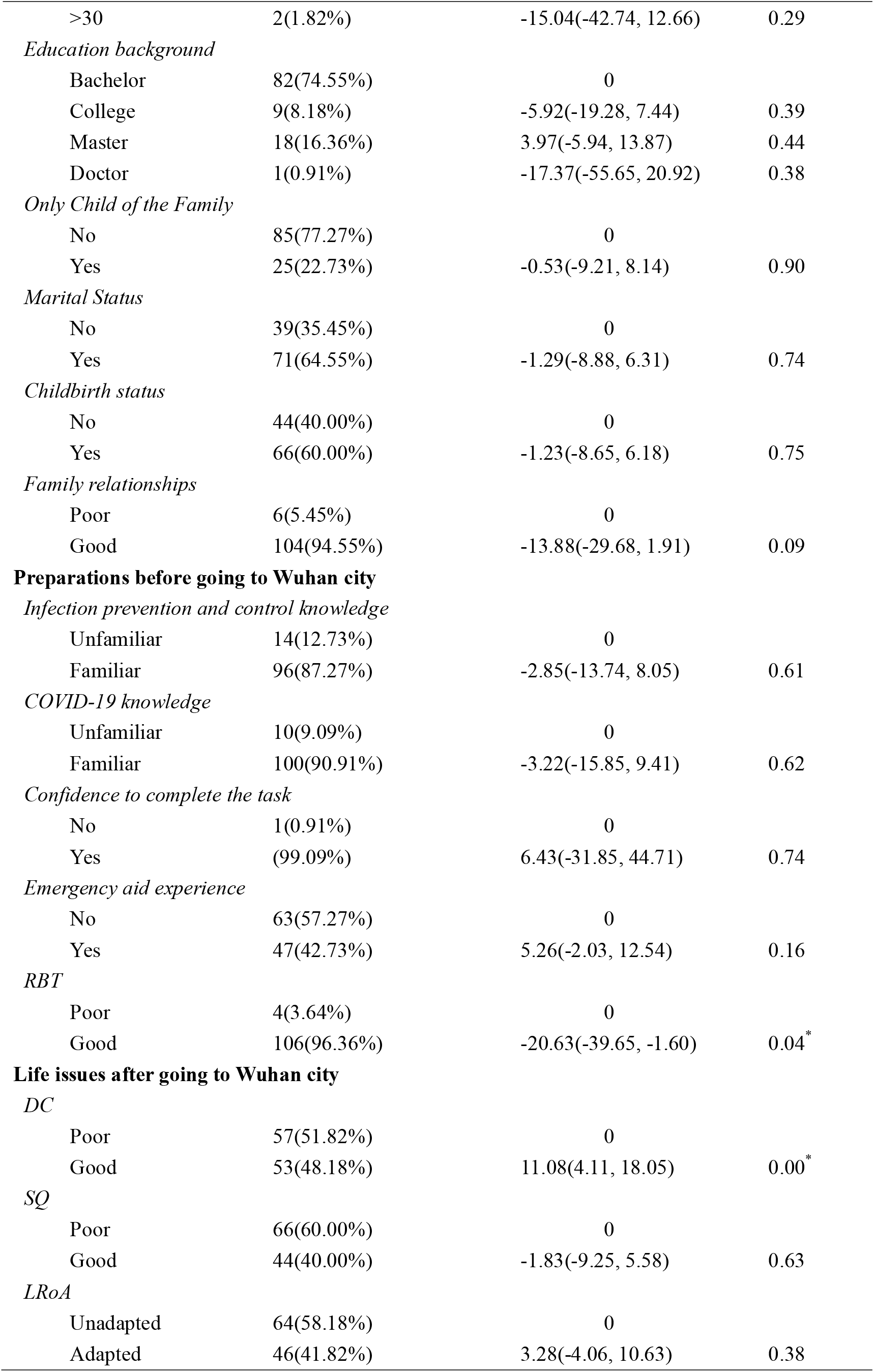
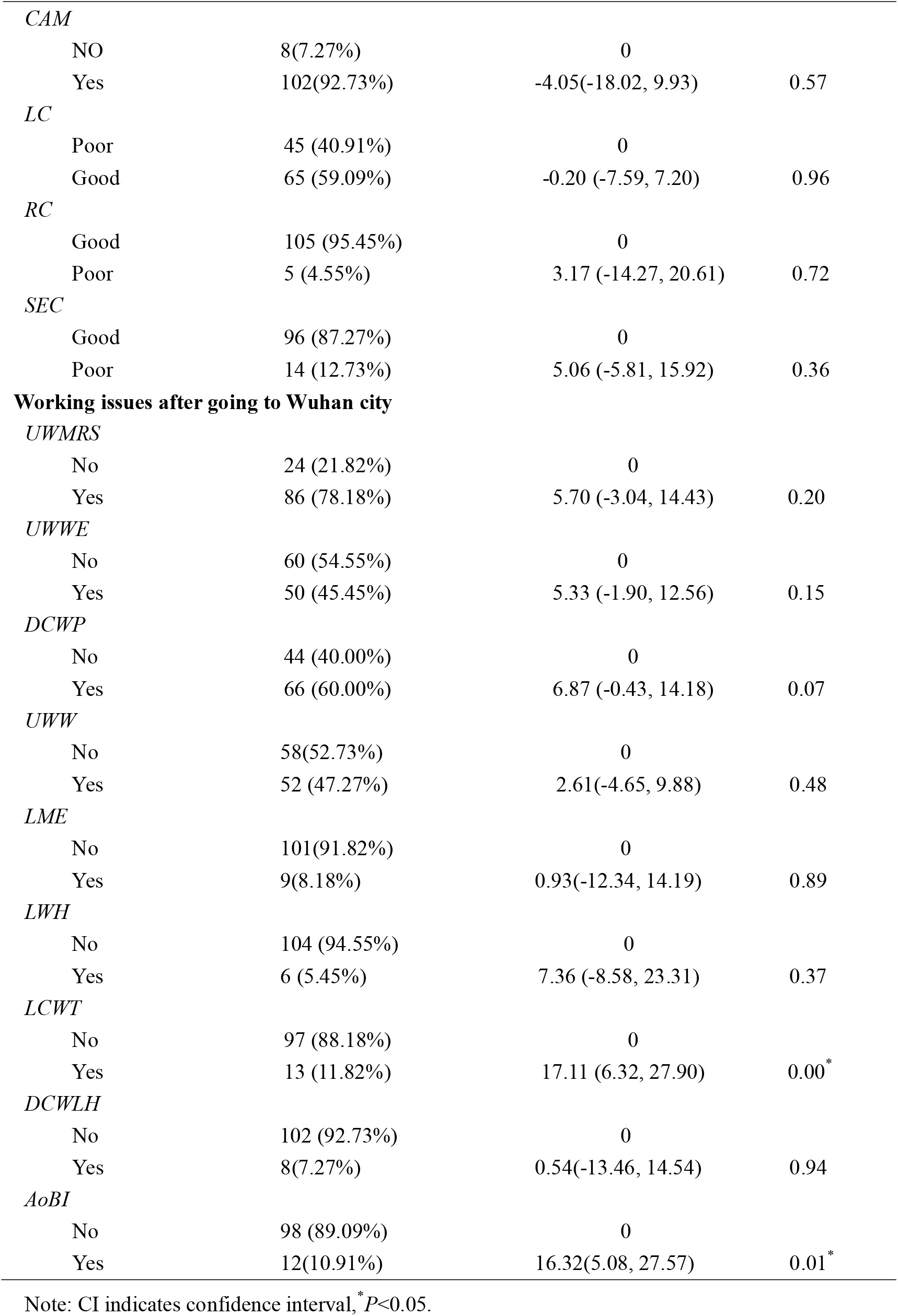
Crude correlation associations of exposure risk factors and GSI of the members

### 3.4 Multivariate logistic regression model for DC, LCWT, AoBI, RBT and GSI score of the members

Multivariate logistic regression analysis showed that, for DC, LCWT, AoBI and RBT, when unadjusted covariates, DC,LCWT and AoBI had a positive correlations with GSI score(*P*<0.05), RBT had a negative correlation with GSI score (*P*<0.05); When adjusted age and gender, DC,LCWT and AoBI also had a positive correlations with GSI score(*P*<0.05), RBT also had a negative correlation with GSI; For DC, when adjusted LCWT, AoBI and RBT, DC also had a positive correlation with GSI score(*P*=0.00<0.05), estimated change was 10.17, 95%CI was 3.30 to 17.04; For LCWT, when adjusted DC, AoBI and RBT, LCWT also had a positive correlation with GSI score(*P*=0.04<0.05), estimated change was 11.55, 95%CI was 0.40 to 22.71; For AoBI, when adjusted DC, LCWT and RBT, AOBI did not had a correlation with GSI score(*P*=0.16>0.05), estimated change was 11.55, 95%CI was 0.40 to 22.71, this result showed that if control other covariates, AOBI will not influence GSI score. For RBT, when adjusted DC, LCWT and AoBI, RBT also had a negative positive correlation with GSI score (*P*=0.00<0.05), estimated change was -28.09, 95%OR was -45.79 to -10.40.(Table3)

**Table3.** Multivariate logistic regression model for DC, LCWT, AoBI, RBT and GSI of the members

| Variables in model | Non-adjusted |  | Adjust I |  | Adjust II |  |
| --- | --- | --- | --- | --- | --- | --- |
| | $\beta$ (95%CI) | *P Value* | $\beta$ (95%CI) | *P Value* | $\beta$ (95%CI) | *P Value* |
| DC |  |  |  |  |  |  |
| No | 0 |  | 0 |  | 0 |  |
| Yes | 11.08(4.11,18.05) | 0.00* | 10.85(3.61,18.09) | 0.00* | 10.17(3.30,17.04) | 0.00* |
| LCWT |  |  |  |  |  |  |
| No | 0 |  | 0 |  | 0 |  |
| Yes | 17.11(6.32,27.90) | 0.00* | 16.54(5.28,27.81) | 0.01* | 11.55(0.40,22.71) | 0.04* |
| AoBI |  |  |  |  |  |  |
| No | 0 |  | 0 |  | 0 |  |
| Yes | 16.32(5.08,27.57) | 0.01* | 15.66(4.11,27.21) | 0.01* | 8.51(-3.14,20.15) | 0.16 |
| RBT |  |  |  |  |  |  |
| Good | 0 |  | 0 |  | 0 |  |
| Poor | -20.63(-39.65,-1.60) | 0.04* | -22.40(-41.89,-2.90) | 0.03* | -28.09(-45.79,-10.40) | 0.00* |

Note: Non-adjusted model adjust for: None; Adjust I model adjust for: age and gender; \* $P < 0.05$ . DC: Adjust II model adjust for: LCWT, AoBI, RBT. LCWT: Adjust II model adjust for: DC, AoBI, RBT. AoBI: Adjust II model adjust for: DC, LCWT, RBT. RBT: Adjust II model adjust for: DC, LCWT, AoBI.

### 3.5 Stratification analysis of DC, LCWT, AoBI, RBT and GSI score of the members between genders

Multivariate logistic regression stratification analysis showed that DC, LCWT and AoBI of female had a positive correlation with GSI score for non- adjusted model and adjusted I model, RBT of female had a negative correlation with GSI score for non- adjusted model and adjusted I model, the differences had statistically significant(*P*<0.05). But for male, above factors had no correlation with GSI score for non-adjusted model, the differences did not had statistically significant (*P*>0.05), DC and RBT had no correlation with GSI score for adjusted I model, the differences also did not had statistically significant (*P*>0.05), LCWT and AoBI had correlation with GSI score for adjusted I model, the differences had statistically significant (*P*<0.05). This results showed that when adjusted age, for female, the relationships of DC, LCWT,AoBI, RBT and GSI score didn’t changed. But for male, when adjusted age, LCWT and AoBI correlations with GSI score started to appear, which showed that age may was an effect modifier. The adjusted II model showed when adjusted covariates of LCWT, AoBI and RBT, DC had a positive correlation with GSI score of female, estimated change was 13.20, 95%CI was 4.55 to 21.85, the difference was statistically significant(*P*=0.00<0.05), but for male was not(*P*=0.59>0.05). RBT had a negative correlation with GSI score of female, when adjusted covariates of DC, LCWT and AoBI, the estimated change was -57.85,95%CI was -94.52 to -21.18, the difference was statistically significant(*P*=0.00<0.05), for male was also not(*P*=0.08>0.05). LCWT and AoBI didn’t had correlation with GSI score between genders, the differences were not statistically significant (*P*>0.05), when adjusted covariates of DC and RBT in adjusted II model (table4).

**Table4.**
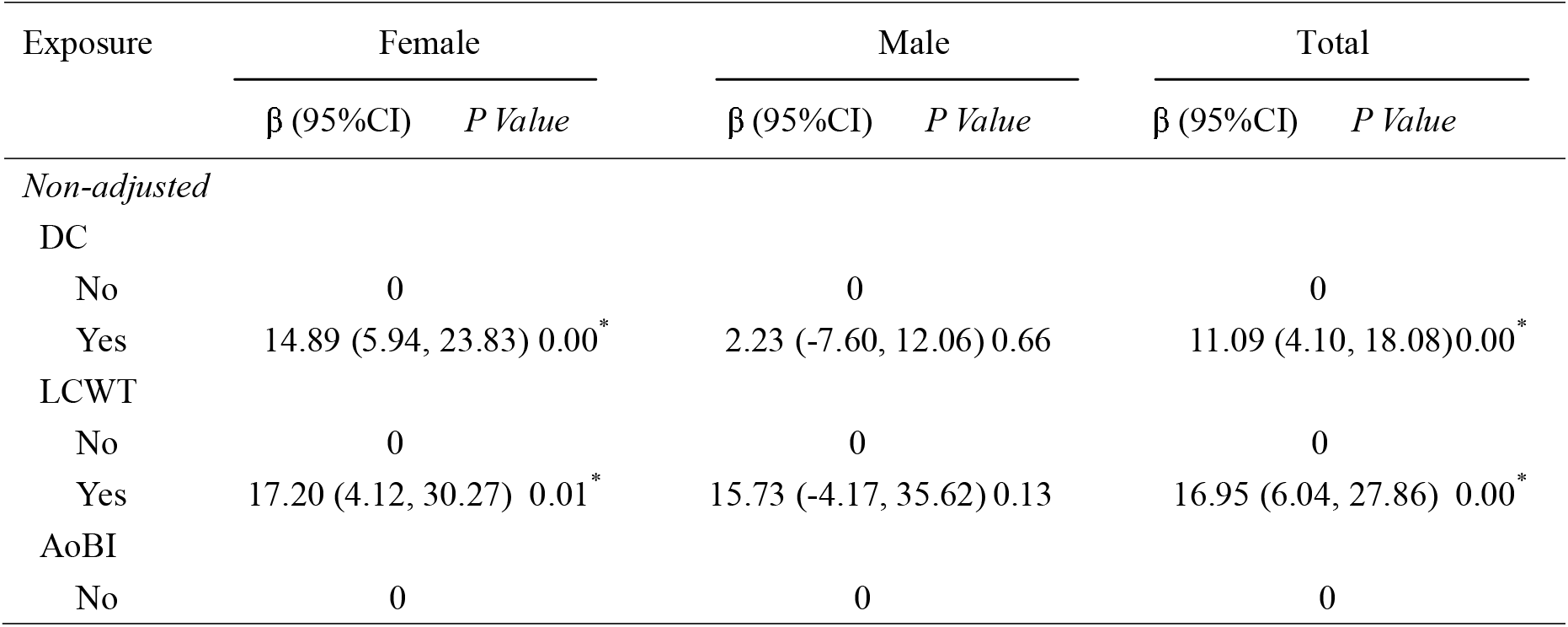
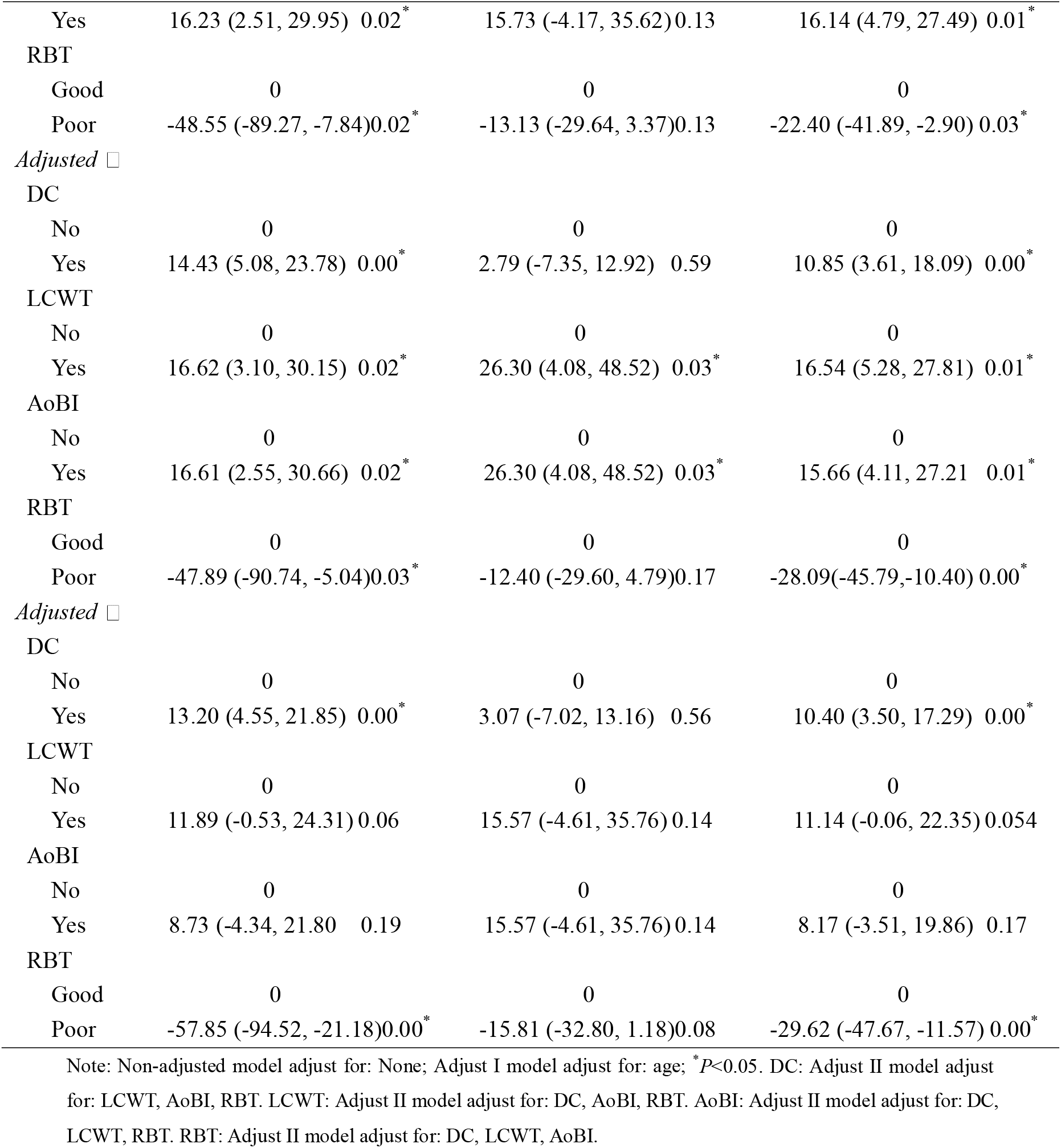
Stratification analysis of DC, LCWT, AoBI, RBT and GSI score of the members between genders

| Exposure | Female |  | Male |  | Total |  |
| --- | --- | --- | --- | --- | --- | --- |
| | $\beta$ (95%CI) | *P Value* | $\beta$ (95%CI) | *P Value* | $\beta$ (95%CI) | *P Value* |
| *Non-adjusted* |  |  |  |  |  |  |
| DC |  |  |  |  |  |  |
| No | 0 |  | 0 |  | 0 |  |
| Yes | 14.89 (5.94, 23.83) | 0.00* | 2.23 (-7.60, 12.06) | 0.66 | 11.09 (4.10, 18.08) | 0.00* |
| LCWT |  |  |  |  |  |  |
| No | 0 |  | 0 |  | 0 |  |
| Yes | 17.20 (4.12, 30.27) | 0.01* | 15.73 (-4.17, 35.62) | 0.13 | 16.95 (6.04, 27.86) | 0.00* |
| AoBI |  |  |  |  |  |  |
| No | 0 |  | 0 |  | 0 |  |

|  |  |  |  |
| --- | --- | --- | --- |
| Yes | 16.23 (2.51, 29.95) 0.02* | 15.73 (-4.17, 35.62)0.13 | 16.14 (4.79, 27.49) 0.01* |
| RBT |  |  |  |
| Good | 0 | 0 | 0 |
| Poor | -48.55 (-89.27, -7.84)0.02* | -13.13 (-29.64, 3.37)0.13 | -22.40 (-41.89, -2.90) 0.03* |
| Adjusted ☐ |  |  |  |
| DC |  |  |  |
| No | 0 | 0 | 0 |
| Yes | 14.43 (5.08, 23.78) 0.00* | 2.79 (-7.35, 12.92) 0.59 | 10.85 (3.61, 18.09) 0.00* |
| LCWT |  |  |  |
| No | 0 | 0 | 0 |
| Yes | 16.62 (3.10, 30.15) 0.02* | 26.30 (4.08, 48.52) 0.03* | 16.54 (5.28, 27.81) 0.01* |
| AoBI |  |  |  |
| No | 0 | 0 | 0 |
| Yes | 16.61 (2.55, 30.66) 0.02* | 26.30 (4.08, 48.52) 0.03* | 15.66 (4.11, 27.21) 0.01* |
| RBT |  |  |  |
| Good | 0 | 0 | 0 |
| Poor | -47.89 (-90.74, -5.04)0.03* | -12.40 (-29.60, 4.79)0.17 | -28.09(-45.79,-10.40) 0.00* |
| Adjusted ☐ |  |  |  |
| DC |  |  |  |
| No | 0 | 0 | 0 |
| Yes | 13.20 (4.55, 21.85) 0.00* | 3.07 (-7.02, 13.16) 0.56 | 10.40 (3.50, 17.29) 0.00* |
| LCWT |  |  |  |
| No | 0 | 0 | 0 |
| Yes | 11.89 (-0.53, 24.31)0.06 | 15.57 (-4.61, 35.76)0.14 | 11.14 (-0.06, 22.35)0.054 |
| AoBI |  |  |  |
| No | 0 | 0 | 0 |
| Yes | 8.73 (-4.34, 21.80) 0.19 | 15.57 (-4.61, 35.76)0.14 | 8.17 (-3.51, 19.86) 0.17 |
| RBT |  |  |  |
| Good | 0 | 0 | 0 |
| Poor | -57.85 (-94.52, -21.18)0.00* | -15.81 (-32.80, 1.18)0.08 | -29.62 (-47.67, -11.57) 0.00* |

Note: Non-adjusted model adjust for: None; Adjust I model adjust for: age; \* $P < 0.05$ . DC: Adjust II model adjust for: LCWT, AoBI, RBT. LCWT: Adjust II model adjust for: DC, AoBI, RBT. AoBI: Adjust II model adjust for: DC, LCWT, RBT. RBT: Adjust II model adjust for: DC, LCWT, AoBI.

## 4. Discussion

The 2019-nCoV(2019 new coronavirus) is officially called SARS-CoV-2 (severe acute respiratory syndrome-coronavirus-2) and the disease is named COVID-19(Corona virus disease-2019), which is currently causing a pandemic in the world. More than 1 million people have been infected with coronavirus, and tens of thousands have died, causing panic in the whole society. People who are not infected are worried and afraid of being infected^7, 14, 15^. Patients who are already infected are worried about whether they can get timely and effective treatment. The pandemic of SARS-CoV-2 has had significant social, psychological and economic consequences worldwide^16^, especially for low-income patients, this is a serious concern when linked to the pandemic^17^.

The outbreak of the SARS-CoV-2 is undesirable by people and needs to be faced with all over the world, but unfortunately, there are too many news reports that SARS-CoV-2 has caused racial hatred and violence in some countries. The occurrence of these incidents is not conducive to fight the epidemic. Medical staffs are the main force to fight against COVID-19. They directly face the patients who have been infected and the chance of being infected is relatively large at work. At present, the psychological research objects of COVID-19 are mainly patients or the general public, but few people pay attention to and study the psychological status of medical staff. A study found that the sudden increase in confirmed cases had brought tremendous stress and anxiety to frontline surgical staff, and researchers believe that early psychological intervention were essential^18^. Researchers from Italy found that at least 2,629 health workers have been infected with coronavirus since the outbreak onset in February, representing 8.3% of total cases^19^. If the medical staffs also feel panic, it may affect their medical behavior, such as avoiding, reducing communication with infected people, or even leaving the job. Therefore, improving the enthusiasm of the medical staff and reducing their panic will play a decisive role in improving the clinical efficacy and even achieving the successful control of the epidemic.

Preliminary evidence suggests that symptoms of anxiety and depression (16-28%) and self-reported stress (8%) are common psychological reactions to the COVID-19 pandemic, and may be associated with disturbed sleep^20^. The outbreak of COVID-19 infection had an effect on the psychology of the elderly people, resulting in anxiety and depression^6^. Our research found that the GSI scores of medical team members were not affected by age, as well as working age, educational background, family relationship, marital status, and fertility status. Before going to Wuhan, medical team members need to receive hospital infection knowledge prevention and control training, learn COVID-19 knowledge, and build confidence in defeating the disease. Knowledge of COVID-19, confidence to complete the task and emergency aid experience also did not have correlations with GSI score. We found that after training, 99.09% members are confident to fight against COVID-19, but just 42.73% members had emergency aid experience and 4 (3.64%) felt that the relationship with other team members was not in harmony. Such research results showed that in order to control the outbreak of COVID-19, China quickly established medical teams, the team members were from various clinical disciplines.

Before going to Wuhan, they did not have time to receive systematic emergency first aid training, and even some team members may not know each other. We have reviewed the literature database in detail, and found no literature about the medical team against COVID-19. A study found that communication had achieved satisfactory results and may play a role in the protection on the psychological condition on the people with close contact with influenza A(H1N1)^21^. Another study showed that patients with difficulty in communicating significant aspects of their complaints to the physician or in understanding his instructions for treatment the authors believed SCL-90 was a quickly administered and easily scored test that can screen for both psychopathology and communication problems^21^. Our experience is worth sharing with the world, but if there is enough time, the members should best to receive systematic emergency first aid training and to increase communication between them, because our research found that RBT and GSI score were negatively correlated, the more harmonious the relationship between the members, the lower the GSI score.

After going to Wuhan, the team members faced life and work issues. Our study showed that DC and AoBI had a positive correlation with GSI score, LCWT had a negative correlation with GSI score for non- adjusted model, but when we adjusted age, gender and covariates, there was no correlation between AoBI and GSI score, but correlations of DC,RBT and LCWT with GSI score didn’t changed. This result showed that the influence of AoBI on GSI score was influenced by the three covariates of DC, RBT and LCWT. A literature published in 1990 found that refined sucrose and caffeine free diet can significantly decline in depression on all depression measures, including SCL-90, Beck Depression Inventory (BDI), and Interpersonal Style Inventory (ISI)^22^.A study found that adding certain substances to food can improve sleep and depression^23^. Our research found that DC was related to GSI scores. Good diet can improve the psychological status of medical team members, but standards for DC are different for different people. This risk factor is greatly influenced by personal subjective attitudes. Our medical team members came from all corners of China. Everyone has different preferences for food. Individuals in one study having poor diets were more likely to suffer from depression than those eating good diets^24^ .There is a Chinese proverb called "difficult to adjust for mouth" .However, we still recommend the logistics support department of medical team, if time and material conditions permit, it would better try to enrich and diversify the catering, and customize the catering according to each person’s different hobbies, so that can alleviate the psychological anxiety and depression.

If the RBT was not good, it will be easy result in LCWT, there was a correlation between the two, and some researchers had already confirmed that lacking communication with others could increase anxiety or depression^25^. As said above, our study also found if the members had bad RBT and LCWT, the GSI Score will increased. Fighting the SARS-CoV-2 epidemic requires joint efforts of medical and nursing members. Due to the short time, members were transferred from various departments to form a team. Many of them did not know each other and understand each other, which will increase their psychological pressure. Some studies have found that recreational activities can improve psychological conditions ^26, 27^, so we suggest that if there is time between the members, they can participate in some collective recreational activities while ensuring protection, which helps increase mutual understanding and can reduce the chance of bad mood.

This study has a shortcoming due to lack of time, we did not follow up the team members. We will observe the changes of their psychological status in the later period, to further understand the long-term effects of RBT, LCWT and AoBI on GSI score.

## Data Availability

The data can be download from the follow link.
Code:54zy

https://pan.baidu.com/s/1C2VOmbw7LJI4mBkChE_GTA

## Declaration of competing interest

The author(s) declared no potential conflicts of interest with respect to the research, authorship, and/or publication of this article.

## Funding source

This research received no specific grant from any funding agency in the public, commercial, or not-for-profit sectors.

## Disclosure statement

No potential conflict of interest was reported by the authors.

## Author contributions

Cheng Wang conducted analyses and drafted the manuscript in consultation with Jinglong Zhang and Yunyun Fang. All authors provided critical revisions and approved a final version of the manuscript prior to publication.

## Acknowledgements

We sincerely thank Jian Tian, Yan Ma, Jun Wang, Shiyang Zhangand Kai Liu for assorting the data, we thank Changcheng Zheng, Yuyou Zhu, Xuhan Zhang, and Hongzhi Ji for devising and issuing the questionnaire. We also like to thank all the members for their cooperation in our study.

## Notes

### Competing Interest Statement

The authors have declared no competing interest.

